# Effective connectivity during faces processing in major depression – distinguishing markers of pathology, risk, and resilience

**DOI:** 10.1101/2021.04.12.21255310

**Authors:** Seda Sacu, Carolin Wackerhagen, Susanne Erk, Nina Romanczuk-Seiferth, Kristina Schwarz, Janina I. Schweiger, Heike Tost, Andreas Meyer-Lindenberg, Andreas Heinz, Adeel Razi, Henrik Walter

**Affiliations:** Berlin School of Mind and Brain, Humboldt Universität zu Berlin, Germany; Division of Mind and Brain Research, Department of Psychiatry and Psychotherapy CCM, Charité - Universitätsmedizin Berlin, corporate member of Freie Universität Berlin, Humboldt-Universität zu Berlin, and Berlin Institute of Health, Berlin, Germany; Department of Psychiatry and Psychotherapy, Central Institute of Mental Health, Mannheim, Germany; Wellcome Centre for Human Neuroimaging, Institute of Neurology, University College London, United Kingdom; Turner Institute for Brain and Mental Health & Monash Biomedical Imaging, Monash University, Australia

**Keywords:** Major depressive disorder, familial risk, resilience, emotional face processing, effective connectivity, fMRI

## Abstract

**Background:** Aberrant brain connectivity during emotional processing, especially within the fronto-limbic pathway, is one of the hallmarks of major depressive disorder (MDD). However, a lack of systematic approaches in previous studies made it difficult to determine whether a specific alteration in brain connectivity reflects a cause, correlate, or effect of the disorder. The current study aimed to investigate neural mechanisms that correspond to disease, risk and resilience in major depression during implicit processing of emotion cues.

**Methods:** Forty-eight patients with MDD, 49 first-degree relatives of patients with MDD and 103 healthy controls performed a face-matching task during functional magnetic resonance imaging. We used dynamic causal modelling to estimate task-dependent effective connectivity at the subject level. Parametric empirical Bayes was then performed to quantify group differences in effective connectivity.

**Results:** Depressive pathology was associated with decreased effective connectivity from the left amygdala and left dorsolateral prefrontal cortex to the right fusiform gyrus, whereas familial risk for depression corresponded to decreased connectivity from the right orbitofrontal cortex to the left insula and from the left orbitofrontal cortex to the right fusiform gyrus. Resilience for depression was related to increased connectivity from the anterior cingulate cortex to the left dorsolateral prefrontal cortex.

**Conclusions:** Our results suggest that the depressive state alters top-down control of higher visual regions during the processing of emotional faces, whereas increased connectivity within the cognitive control network promotes resilience to depression.

## Introduction

Major depressive disorder (MDD) is a common and debilitating mental health problem with a well-known familial association (1). Due to the contribution of both genetic and environmental factors, individuals with a family history of MDD are at a greater risk for developing depression compared to those without a family history of MDD (2-5). However, the pathway from familial risk to clinical symptoms has not been fully understood in MDD.

Through the elevated risk of developing depression, unaffected first-degree relatives are assumed to share some biological and psychological features with MDD patients (6-7). Altered fronto-limbic connectivity is one of these putative risk markers which is associated with MDD and present independently from clinical status (8). Several studies showed that both depressed patients (9-16) and individuals with a family history of depression (17-19) exhibited decreased connectivity in the fronto-limbic pathway during the processing of facial emotion. Although these results support that abnormal fronto-limbic connectivity can be a risk marker for MDD, previous studies compared either MDD patients or individuals at familial risk for MDD with healthy controls. Therefore, it is not clear to what extent alterations in the fronto-limbic pathways are shared by patients and individuals at familial risk for MDD. A systematic approach is, thus, necessary to dissociate disease-specific and risk-related alterations in this circuitry.

On the other hand, despite the familial risk for psychiatric disorders, which is comprised of both genetic and environmental factors, many first-degree relatives do not develop depression and stay psychologically healthy. This phenomenon can be described as resilience, i.e. the maintenance of mental health despite adversity (20-21). It is possible that neural or neurodevelopmental mechanisms in healthy relatives enable this resilience capacity, and that these are unique to the relatives group compared to both patients and controls. Given the high prevalence rate and poor treatment outcomes for MDD (22-23), identifying biopsychosocial factors promoting resilience, as well as the neural mechanism underlying it, becomes important to develop novel approaches to the prevention and treatment of depression (24).

Although research into the neuroscience of resilience is relatively new, previous studies have suggested that brain regions involved in cognitive control of emotions are associated with resilience to depression (21,25). One of these brain regions associated with resilience capacity is the anterior cingulate cortex (24). Previous neuroimaging studies showed that resilience to depression is linked to greater anterior cingulate cortex volume (25-26), greater anterior cingulate cortex activation during a cognitive task (27) and enhanced connectivity between anterior cingulate cortex and the prefrontal cortex during the processing of emotional faces (19,28). Taken together, these findings suggest that the connectivity of the anterior cingulate cortex may be a promising target to test resilience mechanism for MDD.

Here, we re-assessed data reported by Wackerhagen *et al*. (28) and adopted the same group-comparative approach (Supplementary Material S1) to disentangle disease, risk and resilience in neuro-functional markers for MDD during an implicit emotion processing task. However, instead of using generalized psychophysiological interactions (29) to assess task-dependent functional connectivity, we utilized Dynamic Causal Modelling (DCM) to measure effective connectivity in the target neural pathways. This brings two main methodological advantages to the present study. First, unlike functional connectivity, effective connectivity allows us to determine the direction of influences (i.e., causal relationships in the context of the model) as well as the valence of the influence (i.e., inhibitory or excitatory signaling) among coupled brain regions (30-31). Second, the majority of task-dependent functional connectivity studies used amygdala as a seed region and investigated the amygdala connectivity with the rest of the brain in MDD patients and individuals at high risk for MDD. Although the amygdala plays a crucial role in depression, we wanted to allow for new insights into the neural model of MDD by utilizing a model-based approach that takes into account the connectivity patterns of other important brain regions, such as the prefrontal cortex (32-33) and high-level visual regions like the fusiform gyrus (34-36).

Based on our previous study (28), we hypothesized that decreased effective connectivity within the fronto-limbic pathway will be identified as disease and/or risk factor for MDD, whereas connectivity between anterior cingulate cortex and prefrontal cortex will be related to resilience to MDD. Additionally, there is a growing literature showing that MDD patients have functional abnormalities in visual cortex regions during emotional face processing (34-40). Therefore, we wanted to explore whether altered effective connectivity of fusiform gyrus will be associated with disease state and/or risk for MDD. Finally, we explored the relationship between negative affect and task-related connectivity to evaluate the functional relevance of potential alterations.

## Methods and Materials

### Participants

Forty-eight patients with MDD (34 females, mean age=31.25), 49 first-degree relatives of patients with MDD (33 females, mean age=28.49) and 103 healthy controls (61 females, mean age=31.88) were selected from two multicenter studies on the neuro-genetic causes of major depression, schizophrenia, and bipolar disorder (28,41). First-degree relatives were included in this study if they had at least one first-degree relative diagnosed with MDD. Except for family history of MDD in first-degree relatives, neither first-degree relatives nor healthy controls had a personal or familial history of lifetime axis I disorders. Inclusion criteria for patients were a current diagnosis of a recurrent depressive disorder or a depressive episode that was either severe or had lasted for at least two years. Psychiatric history was confirmed using the German version of the Structured Clinical Interview for DSM-IV-TR Axis I Disorders (42). Thirty-three of the patients were receiving psychotropic medication at the time of the fMRI investigation (Supplementary Material Table S1). For further details of the study cohorts please see Wackerhagen *et al*. (28).

The study was approved by local ethics committees of the study sites. All participants provided written informed consent.

### Psychological Measurements

The current level of negative affectivity was assessed using the Beck Depression Inventory (43), the depression scale of the Symptom Checklist-90 (SCL-90-R;44), State-Trait Anxiety Inventory-State (STAI-S;45), State-Trait Anxiety Inventory-Trait (STAI-T;45), and the neuroticism scale of the NEO-Five Factor Inventory (NEO-FFI;46). A composite score of negative affect was computed for each participant based on BDI, SCL-90 Depression, STAI-T and NEO-FFI Neuroticism scales using principal component analysis (see Supplementary Material S3 for details of principal component analysis).

### Experimental Paradigm

We used a block-designed fMRI task adapted from Hariri *et al*. (47) to investigate the neural correlates of implicit emotion processing. The task had two conditions: face-matching and shape-matching (Supplementary Material Figure S2). During the face-matching condition, participants were asked to match one of two simultaneously presented faces (angry or fearful faces) with the identical target face. During the sensorimotor control condition, participants matched geometric shapes similarly. Eight blocks (4 blocks for each condition) were presented in alternating order. Each block consisted of 6 trials of 5 seconds and started with a brief instruction. Total task duration was 256 seconds.

### Image Acquisition and Preprocessing

The images were acquired on a Siemens Magnetom Trio (Siemens, Erlangen, Germany) 3T MRI scanner using identical scanning protocols. During the task, 135 functional images were obtained using an asymmetric gradient echo-planar sequence sensitive to blood oxygen level-dependent (BOLD) contrast (28 slices, TE= 30 ms, TR = 2000 ms, flip angle = 80°, FoV = 192 mm, voxel size = 3 × 3 × 4 mm).

Image preprocessing was performed using SPM 12 (http://www.fil.ion.ucl.ac.uk/spm/) and included slice timing correction, motion correction, structural and functional image co-registration, segmentation, normalization to the Montreal Neurological Institute (MNI) 152 template, and smoothing using a kernel with a full-width half-maximum of 8 mm. Furthermore, to quantify mean head motion for each participant, we computed frame-wise displacement based on rigid body transformation parameters (48,49). For a detailed description of image acquisition and preprocessing, see Supplementary Material S5.

### Generalized Linear Modeling

Generalized Linear Modeling (GLM) implemented in SPM12 was performed to estimate brain responses. Each experimental condition (face-matching and shape-matching) and instructions were convolved with a canonical hemodynamic response function. Six motion parameters and time series from WM and CSF were entered into the subject-level analysis as nuisance covariates to correct for motion and physiological noise.

At the group level, a one-sample t-test was performed to find brain regions showing the main effect of task (faces > shapes). Additionally, we conducted one-way ANOVA to examine group differences in the main effect of task. Age, sex, education, study site, and mean head motion were included in all analyses as covariates.

### Regions of Interest Selection

Based on our GLM results, we selected the following regions of interests (ROIs) to use in subsequent DCM analysis: fusiform gyrus, amygdala, anterior cingulate cortex, dorsolateral prefrontal cortex, orbitofrontal cortex, and insula. All ROIs were chosen bilaterally, except the anterior cingulate cortex. The model included 11 ROIs in total. The MNI coordinates of the ROIs can be seen in Table 1.

**Table 1.** Locations of Regions of Interest.

| **Regions of Interest** | **MNI Coordinates** | **T** |
| --- | --- | --- |
| Fusiform Gyrus L | -36 -73 -13 | 29.69 |
| Fusiform Gyrus R | 39 -55 -16 | 28.29 |
| Amygdala L | -24 -4 -19 | 13.88 |
| Amygdala R | 27 -4 -19 | 14.55 |
| Anterior Cingulate Cortex | 9 26 20 | 7.00 |
| Dorsolateral Prefrontal Cortex L | -51 29 23 | 13.39 |
| Dorsolateral Prefrontal Cortex R | 54 32 20 | 17.59 |
| Orbitofrontal Cortex L | -36 32 -16 | 12.19 |
| Orbitofrontal Cortex R | 33 32 -16 | 14.73 |
| Insula L | -36 23 -1 | 8.10 |
| Insula R | 36 29 -1 | 9.00 |

Regions of Interest were located at group-level peak MNI coordinates of the faces > shapes contrast at a significance threshold of $p < 0.05$ (FWE-corrected).

To account for individual differences in the peak locations of brain activation, we searched for the local maxima nearest to the group-level coordinates within anatomical boundaries of given ROIs (thresholded at p < 0.05, uncorrected). Regional responses were then summarized with the first-eigenvariate of all activated voxels within a 6 mm sphere of the subject-specific local maxima. For participants showing no experimental effect within a given ROI, the first eigenvariate of time series was extracted from a 6 mm sphere of the group-level maximum (50). Participants who did not show consistent experimental effect for more than two ROIs within a hemisphere were excluded from further analysis (n=13). For a detailed description, see Supplementary Material S6.

### Dynamic Causal Modelling

Effective connectivity was investigated using DCM for fMRI. DCM is a Bayesian framework which combines a biologically realistic neuronal model with a biophysically validated hemodynamic model to estimate neural responses from the observed fMRI signal. It models intrinsic connections within and between brain regions and modulation of these intrinsic connections by context-dependent inputs (For a detailed description, see Supplementary Material S7).

In this study, we wanted to investigate the effect of viewing emotional faces on intrinsic connections. For this purpose, we specified a fully connected model with 121 neural coupling parameters (see Supplementary Material Figure S5), which allowed us to compare all possible nested models within the network. The driving input (i.e., emotional faces) entered the model through bilateral fusiform gyri and propagated through the network via intrinsic connections. After the model estimation, we performed diagnostics to check the quality of the DCM model fitting to ensure that model inversion was successful (51). Participants whose explained variance by the model was less than 10% (n=8) were excluded from further analyses. The final sample for group-level DCM analyses included 179 participants (see Supplementary Material Figure S6 for the flow diagram of study participants).

### Empirical Bayes for Group DCM

We quantified commonalities and group differences in effective connectivity using Parametric Empirical Bayes (PEB). The PEB is a hierarchical Bayesian framework to estimate effective connectivity parameters at the group level. Here, we set three PEB analyses to compare three groups (e.g., controls versus patients, controls versus relatives and relatives versus patients). Age, sex, education, study site and mean head motion were included in all analyses as covariates (For a detailed description see Supplementary Material S9). Since PEB is a multivariate Bayesian GLM, in which all the connectivity parameters are fitted at once to optimize the model evidence, no correction for multiple comparisons is required in contrast to a frequentist approach.

After group differences in effective connectivity were determined, we used a similar framework as in Wackerhagen *et al*. (28) to associate these group differences with disease pathology, risk and resilience. For disease-related changes in effective connectivity, we looked at both ‘controls versus patients’ and ‘relatives versus patients’ contrasts and determined the connections which are altered in patients compared to both controls and relatives. In the same way, we looked at both ‘controls versus patients’ and ‘controls versus relatives’ contrasts to determine shared features by patients and relatives (i.e., risk-related changes). Finally, we looked at ‘controls versus relatives’ and ‘relatives versus patients’ contrasts and determined the connections which are altered in relatives compared to both patients and controls to identify resilience-related changes in effective connectivity.

In addition, we explored the associations between effective connectivity strengths and negative affect scores for each group separately using the PEB framework.

## Results

### Behavioral Results

As shown in Table 2, groups did not differ in terms of age, sex, years of education and task performance. However, there were significant group differences in study site and head motion. Patients had significantly higher head motion (Mdn = 0.12) than controls (Mdn = 0.09, p = 0.04) and relatives (Mdn = 0.08, p< 0.001). Moreover, as expected, the one-way ANOVA test revealed a significant main effect of group in all psychological measurements (For test statistics of post-hoc comparisons, see Supplementary Material Table S4).

**Table 2.** Sample Characteristics.

|  | **Controls**<br>**(N=103)** | **Relatives**<br>**(N=49)** | **Patients**<br>**(N=48)** | **df** | **F $\chi^2$** | **p** |
| --- | --- | --- | --- | --- | --- | --- |
| **Demographics** |  |  |  |  |  |  |
| Sex, M/F, N | 42/61 | 16/33 | 14/34 | 2 | 2.23 | 0.33 |
| Age (y), M (SD) | 31.25 (9.33) | 28.49 (8.11) | 31.88 (8.97) | 197 | 2.10 | 0.12 |
| Education (y), M (SD) | 14.14 (2.89) | 14.49 (2.80) | 14.78 (2.19) | 2 | 0.92 | 0.40 |
| Study Site, N |  |  |  | 4 | 26.76 | <0.001<sup>a</sup> |
| Bonn | 13 | 16 | 0 |  |  |  |
| Mannheim | 30 | 15 | 25 |  |  |  |
| Berlin | 60 | 18 | 23 |  |  |  |
| Mean Head Motion, Mdn (IQR) | 0.09 (0.05) | 0.08 (0.04) | 0.12 (0.10) | 197 | 12.67 | 0.002<sup>a</sup> |
| Type of Familial Relationship,<br>N |  |  |  |  |  |  |
| Offspring/Sibling/Parent |  | 35/12/1 |  |  |  |  |
| **Task Performance** |  |  |  |  |  |  |
| Reaction Time, sec, M (SD) |  |  |  |  |  |  |
| Shapes | 1.11 (0.21) | 1.12 (0.29) | 1.12 (0.19) | 197 | 0.02 | 0.98 |
| Faces | 1.21 (0.24) | 1.25 (0.30) | 1.19 (0.19) | 197 | 0.74 | 0.48 |
| Accuracy, n (%) |  |  |  |  |  |  |
| Shapes | 23.18(96.58) | 23.32(97.21) | 23.27(96.96) | 196 | 0.22 | 0.80 |
| Faces | 23.64 (98.5) | 23.67(98.62) | 23.75(98.96) | 196 | 0.19 | 0.83 |
| **Psychological Measurements** |  |  |  |  |  |  |
| SCL-90 Depression, Mdn (IQR) | 0.08 (0.29) | 0.08 (0.27) | 1.54 (1.27) | 197 | 88.26 | <0.001<sup>a</sup> |
| BDI, Mdn (IQR) | 1 (3.5) | 3 (4.5) | 21 (16.5) | 197 | 99.78 | <0.001<sup>a</sup> |
| STAI-S, M (SD) | 31.56 (6.51) | 31.85 (5.32) | 49.72(10.89) | 162 | 89.88 | <0.001<sup>a</sup> |
| STAI-T, M (SD) | 33.46 (8.43) | 35.76 (8.10) | 56.53(10.65) | 191 | 112.32 | <0.001<sup>a</sup> |
| NEO-FFI Neuroticism, M (SD) | 14.32 (6.62) | 17.5 (6.99) | 32.53 (8.04) | 193 | 110.80 | <0.001<sup>a</sup> |
| Negative Affect, Mdn (IQR) | -0.56 (0.60) | -0.35 (0.73) | 1.59 (0.98) | 191 | 52.95 | <0.001<sup>a</sup> |

<sup>a</sup> p < 0.05
Abbreviations: BDI, Beck's Depression Inventory; HDRS, Hamilton Depression Rating Scale; NEO, NEO-Five Factor Inventory; SCL90-R Depression, Symptom Checklist 90 Revised Depression Scale; STAI-S, State Trait Anxiety Inventory - State Anxiety; STAI-T, State Trait Anxiety Inventory - Trait Anxiety

### Task-Related Brain Activity

Across participants, the face-matching task elicited more activation than the shape-matching task in the visual cortex, fusiform gyrus, dorsal prefrontal cortex, subcortical areas (thalamus, amygdala, hippocampus, putamen) and cerebellum (p < 0.05, whole-brain FWE corrected; Figure 1 and Supplementary Material Table S5). The shape-matching task elicited more activation than the face-matching task in bilateral parietal lobes, middle and anterior cingulate cortex, middle occipital cortex, and middle frontal gyrus (p < 0.05, whole-brain FWE corrected).

**Figure 1.**
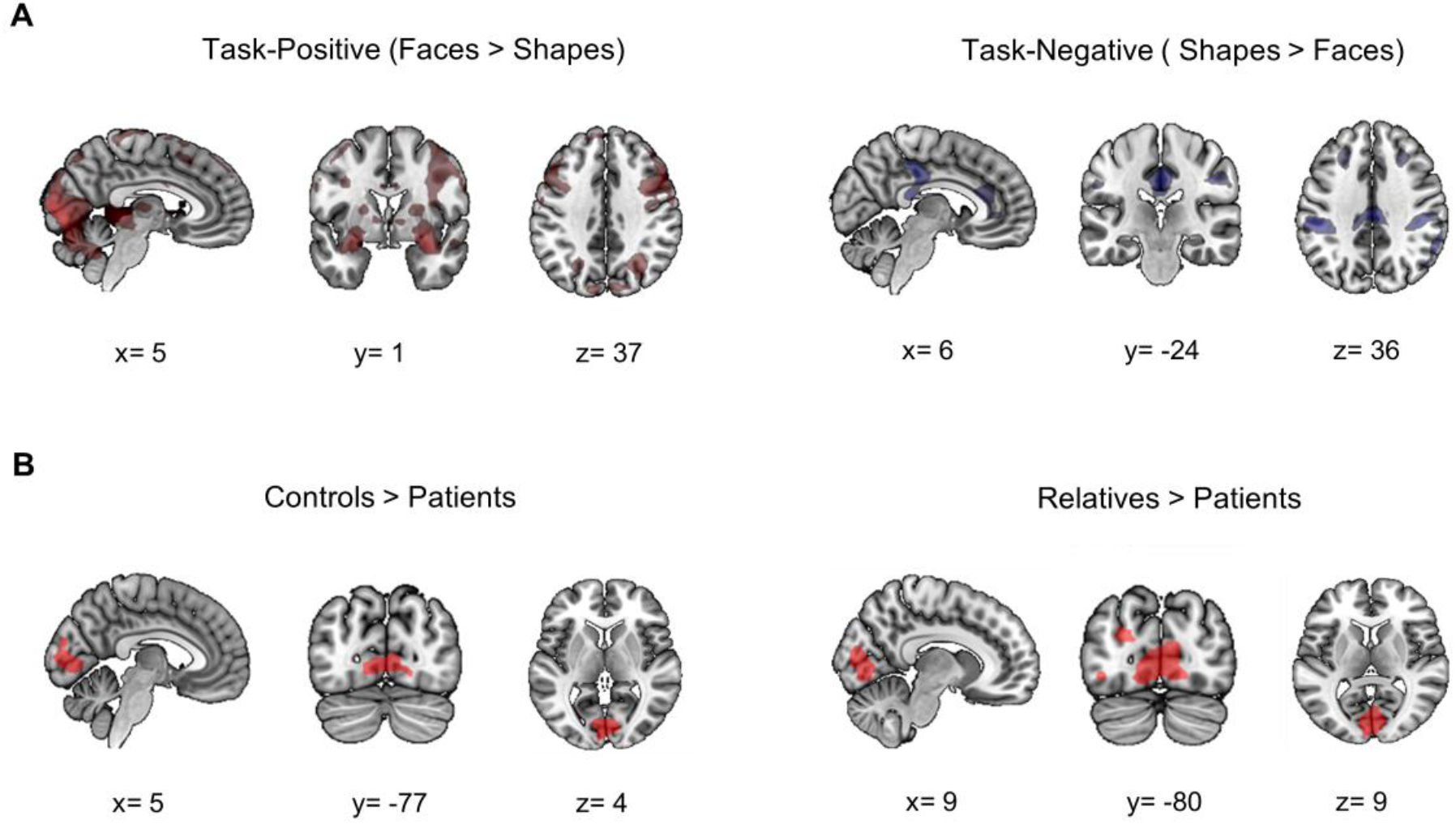
Task-related brain activity across and between groups. The upper panel (A) shows brain regions exhibiting increased (left panel, red color) and decreased (right panel, blue color) activation during the face-matching condition compared to the shape-matching condition (p < 0.05; whole-brain FWE corrected). The lower panel (B) shows group differences in the task-related activation in healthy controls (left panel) and first-degree relatives (right panel) compared to patients with MDD. Here, red color indicates increased regional responses during the face-matching condition relative to the shape-matching condition in the respective group comparison.

The one-way ANOVA test showed a significant effect of group on brain responses (faces > shapes). Post-hoc group comparisons revealed that patients exhibited significantly decreased activation in visual cortex (bilateral lingual gyrus, superior occipital gyrus and calcarine sulcus) compared to controls and relatives (p < 0.05, whole-brain FWE corrected; Figure 1 and Supplementary Material Table S6. We did not observe any significant group difference between controls and relatives in task-related brain activation (p < 0.05, whole-brain FWE corrected).

### Effective Connectivity

Between-group differences in effective connectivity during faces-matching condition are listed in Table 3. We here report only the connection parameters with a probability greater than 95% (posterior probability > 0.95), which corresponds to strong evidence. Having determined between-group differences, we identified connection parameters which were associated with depressive state, risk and resilience for MDD.

**Table 3.** Between-group differences of effective connectivity during the face-matching task.

|  | **Connection**<br>**Parameter** | **Effect size**<br>**(Posterior**<br>**Expectation)** | **95% Bayesian**<br>**Confidence**<br>**Interval** |
| --- | --- | --- | --- |
| **Controls > Patients** |  |  |  |
|  | AMG R → FG R | 0.04 | [0.01, 0.06] |
|  | AMG R → FG L | 0.03 | [-0.001, 0.05] |
|  | OFC R → INS L | 0.03 | [0.01, 0.05] |
|  | AMG L → FG R | 0.07 | [0.04, 0.10] |
|  | OFC L → FG R | 0.07 | [0.05, 0.10] |
|  | OFC L → FG L | 0.08 | [0.05, 0.12] |
|  | dIPFC L → FG R | 0.09 | [0.06, 0.12] |
|  | dIPFC L → OFC R | 0.04 | [0.01, 0.06] |
|  | dIPFC L → dIPFC R | 0.03 | [0.00, 0.06] |
|  | dIPFC L → FG L | 0.09 | [0.06, 0.12] |
|  | dIPFC L → AMG L | 0.03 | [-0.001, 0.06] |
|  | dIPFC L → INS L | 0.02 | [-0.01, 0.05] |
| **Controls < Patients** |  |  |  |
|  | dIPFC R → FG R | -0.02 | [-0.05, 0.001] |
|  | AMG L → INS L | -0.02 | [-0.05, -0.000] |
| **Relatives > Patients** |  |  |  |
|  | ACC → dIPFC L | 0.03 | [0.01, 0.05] |
|  | ACC → INS L | 0.02 | [0.000, 0.03] |
|  | AMG L → FG R | 0.06 | [0.03, 0.09] |
|  | OFC L → FG L | 0.05 | [0.02, 0.08] |
|  | dIPFC L → FG R | 0.08 | [0.05, 0.12] |
|  | INS L → FG R | 0.07 | [0.04, 0.10] |
|  | INS L → OFC L | 0.05 | [0.02, 0.08] |
| **Controls > Relatives** |  |  |  |
|  | OFC R → FG L | 0.04 | [0.02, 0.06] |
|  | OFC R → INS L | 0.02 | [0.00, 0.04] |
|  | OFC L → FG R | 0.04 | [0.01, 0.06] |
| **Controls < Relatives** |  |  |  |
|  | ACC → AMG L | -0.01 | [-0.02, 0.001] |
|  | ACC → dIPFC L | -0.02 | [-0.04, -0.003] |
|  | FG L → FG R | -0.03 | [-0.05, -0.01] |

Abbreviations. ACC, anterior cingulate cortex; AMG, amygdala; dIPFC, dorsolateral prefrontal cortex;
FG, fusiform gyrus; INS, insula; L, left; OFC, orbitofrontal cortex; R, right.

Disease state was associated with lower effective connectivity from left amygdala and left dorsolateral prefrontal cortex to right fusiform gyrus and from left orbitofrontal cortex to left fusiform gyrus (Figure 2). As seen in Figure 2, the estimated group means for these intrinsic connection parameters had negative values. That is, high activity in the source region leads to a decrease in activity in the target region (i.e., inhibitory influence). Thus, lower effective connectivity observed in MDD patients corresponds to more inhibitory influence from amygdala and frontal regions to fusiform gyrus. Furthermore, analysis of brain-behavior relationships revealed that effective connectivity from left amygdala to right fusiform gyrus was negatively associated with negative affect scores in MDD patients (standardized β coefficient = -0.08, posterior probability > 0.95), whereas effective connectivity from left dorsolateral prefrontal cortex to right fusiform gyrus was positively associated with negative affect scores in MDD patients (standardized β coefficient = 0.08, posterior probability > 0.95).

**Figure 2.**
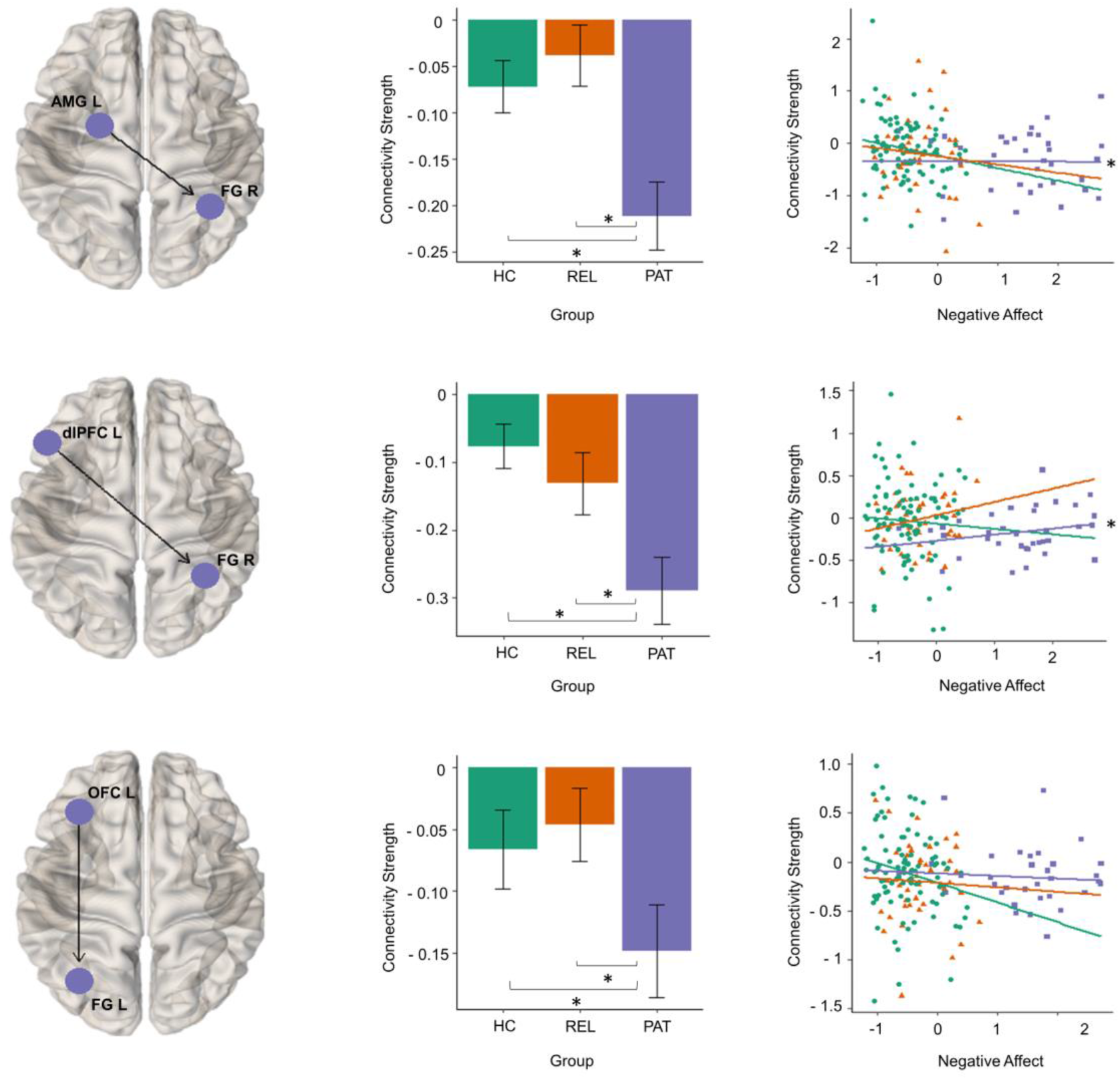
Disease-related alterations in effective connectivity. The middle panel shows the group means of the effective connection parameters which were associated with depressive pathology. Error bars indicate 95% Bayesian confidence interval. Group differences with strong evidence (i.e., posterior probability (free energy with versus without parameter) are larger than .95) are marked with an asterisk. Negative affect scores and estimated posterior means of connection parameters are plotted in the right panel for visualization. The associations with strong evidence are marked with an asterisk. Abbreviations. HC, healthy controls; PAT, patients with major depression; REL, first-degree relatives of patients with depression.

Risk for MDD was associated with decreased effective connectivity from right orbitofrontal cortex to left insula and from the left orbitofrontal cortex to right fusiform gyrus (Figure 3). Controls exhibited higher effective connectivity in these connection parameters compared to relatives and patients. In addition, the alteration in effective connectivity from left orbitofrontal cortex to right fusiform gyrus was gradual (patients < relatives < controls) and negatively associated with negative affect scores in MDD patients (standardized β coefficient = -0.11, posterior probability > 0.95).

**Figure 3.**
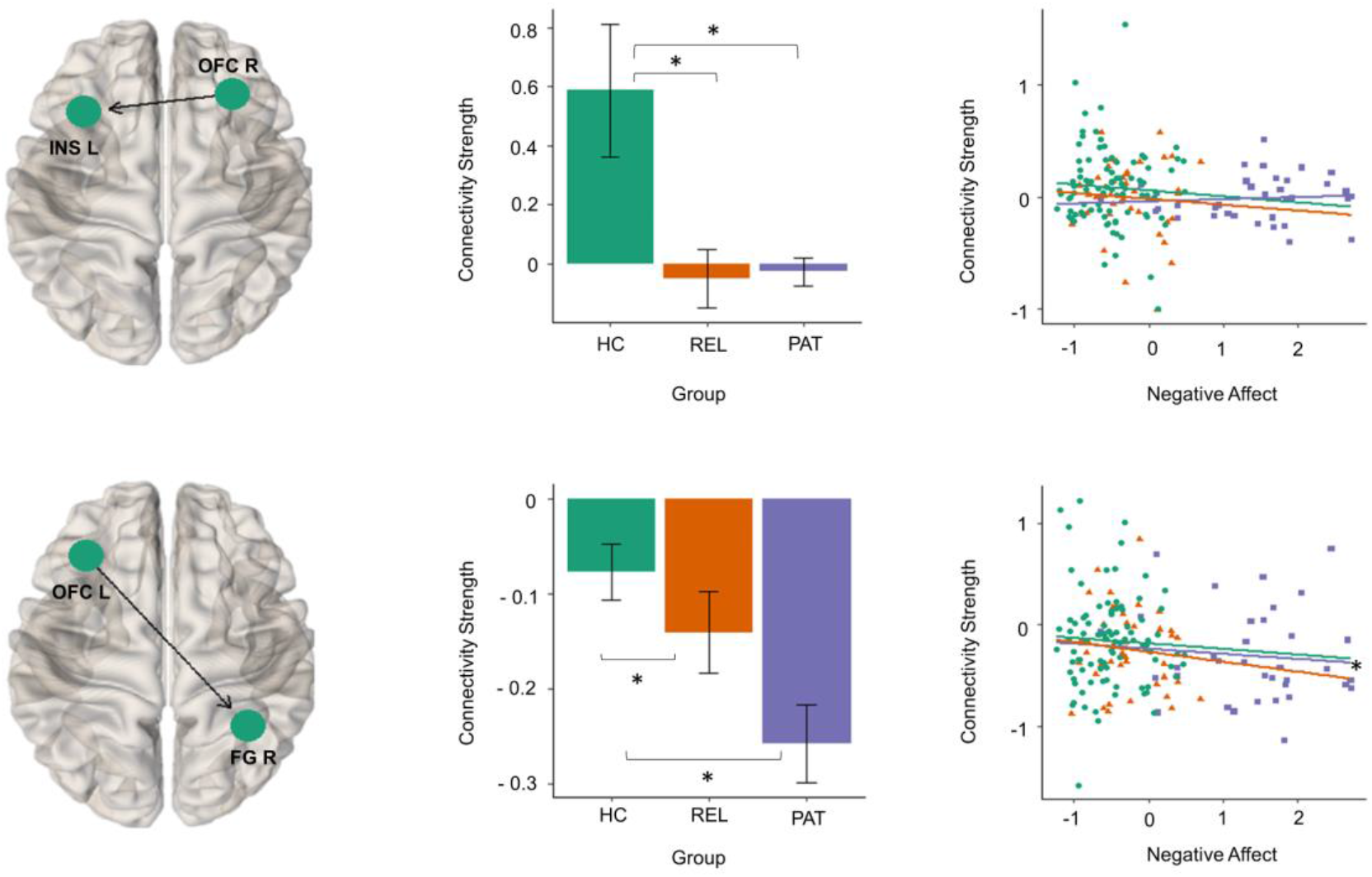
Risk-related alterations in effective connectivity. The middle panel shows the group means of the effective connection parameters which were associated with risk for depression. Error bars indicate a 95% Bayesian confidence interval. Group differences with strong evidence are marked with an asterisk. Negative affect scores and estimated posterior means of connection parameters are plotted in the right panel for visualization. Associations with strong evidence are marked with an asterisk. Abbreviations. HC, healthy controls; PAT, patients with major depression; REL, first-degree relatives of patients with depression.

Effective connectivity from anterior cingulate cortex to left dorsolateral prefrontal cortex was elevated in relatives compared to controls and patients in the resilience contrast (Figure 4). A positive association between negative affect scores and connectivity strength was detected in MDD patients (standardized β coefficient = 0.13, posterior probability > 0.95).

**Figure 4.**
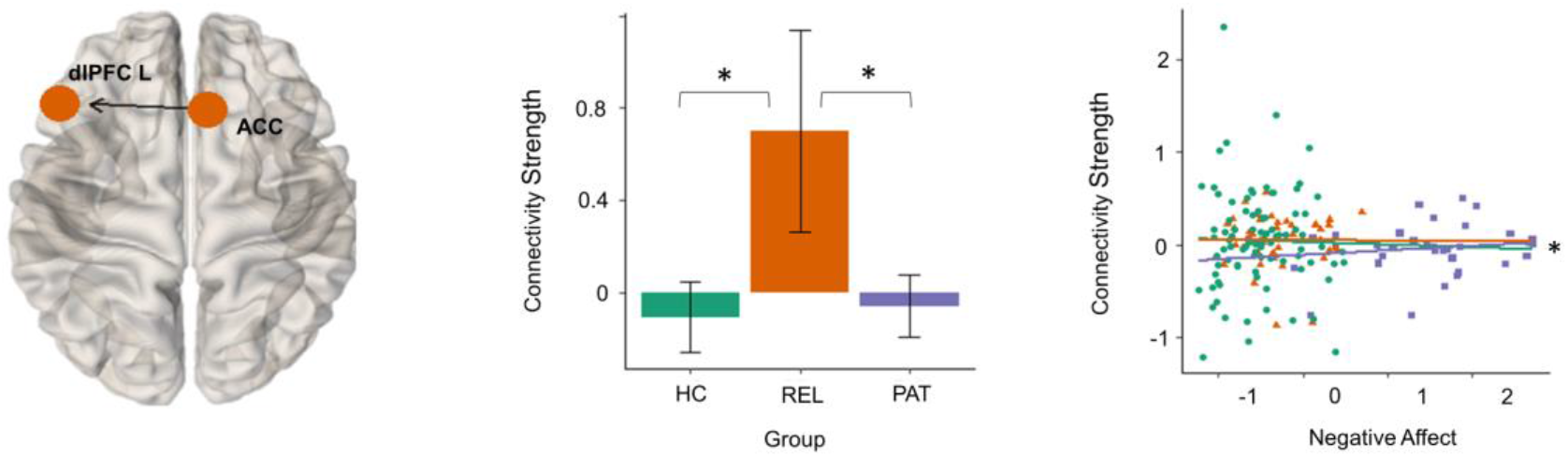
Resilience-related alterations in effective connectivity. The middle panel shows the group means of the effective connection parameter which was associated with resilience. Error bars indicate 95% Bayesian confidence interval. Group differences with strong evidence are marked with asterisk. Negative affect scores and estimated posterior means of connection parameters are plotted in the right panel for visualization. Associations with strong evidence are marked with an asterisk. Abbreviations. HC, healthy controls; PAT, patients with major depression; REL, first-degree relatives of patients with depression.

## Discussion

The present study investigated brain responses and effective connectivity during the face-matching task in patients with MDD, unaffected first-degree relatives and healthy participants. Our results revealed that the groups differed in task-related activity and effective connectivity in the absence of group differences in task performance.

### Disease State

Compared to relatives and controls, MDD patients exhibited lower activity in several visual areas and lower effective connectivity (i.e., more inhibitory influence) from higher order areas to fusiform gyrus during the face-matching task. Specifically, MDD patients showed more inhibitory influence from the left amygdala to right fusiform gyrus than controls and relatives. This increased inhibitory influence was also associated with higher negative affect in MDD patients. Several studies showed that the amygdala has a modulatory role over the visual cortex during emotional face processing (52-57). It can enhance or diminish sensory representation of a stimulus in visual regions based on its motivational significance (58). More inhibitory influence in patients with higher negative affect indicates that this alteration could arise due to an increased perception of negative emotion (59). The fusiform gyrus has a crucial role in face processing and social perception (60-64), therefore, excessive inhibition of fusiform gyrus activity by amygdala can cause diminished visual attention to motivationally important stimuli in MDD patients.

Additionally, disease pathology was associated with increased inhibitory influence from the lateral prefrontal regions to fusiform gyrus. MDD patients with higher negative affect also exerted less inhibitory influence from left dorsolateral prefrontal cortex to right fusiform gyrus. These results are compatible with previous studies reporting decreased functional connectivity between frontal and visual regions in MDD patients during an emotional task (65-66) and at resting-state (67-69). The lateral prefrontal cortex plays an important role in the integration of cognitive and emotional information (70-71) and inhibition of task-irrelevant stimuli (72-74). Moreover, similar to the amygdala, it can modulate the activation of sensory and association cortices (75-76). Previous studies showed that emotional stimulus can interfere goal-directed behavior (72), and top-down control of emotional distractors can result in better task performance (77-78). Therefore, more inhibitory influence detected in patients could be related to inhibition of emotional distractors (e.g., facial expression) during the identity matching and contribute to maintaining the same level of task performance with controls and relatives by preventing the disruptive effect of emotional distractors on working memory (77). However, due to the lack of variability in task performance, we could not test this hypothesis. Moreover, the utilized task is not cognitively challenging. Therefore, the alleged compensatory mechanism cannot be generalized to cognitively demanding tasks since MDD patients may fail to inhibit emotional distractors when task difficulty increases.

### Risk

Decreased effective connectivity from right orbitofrontal cortex to the left insula was associated with risk for MDD. The insula has been related to affective processing and has bidirectional connections with orbitofrontal cortex (79). The increased coupling between the insula and prefrontal cortex in healthy controls was linked to attenuated distraction (80) and decreased negative emotion during suppression (81). Thus, the increased fronto-insular effective connectivity in controls may reflect a functional mechanism, possibly suppression of irrelevant information, which helps participants to deal with emotional distractors, whereas decreased effective connectivity in patients and relatives might reflect increased attention to negative facial expressions and lower suppression success.

Furthermore, similar to MDD patients, relatives exhibited decreased connectivity from left orbitofrontal cortex to right fusiform gyrus compared to controls. A recent study reported that the main effect of familial risk for depression (i.e., patients and healthy controls with versus without family history of MDD) was associated with altered functional connectivity between the orbitofrontal cortex and visual regions (82). Although we did not directly investigate the main effect of familial risk in this study in the same way, our results indicate that orbitofrontal cortex might be an important brain region to investigate risk-related changes in depression.

Contrary to our hypothesis, we did not identify decreased effective connectivity between the amygdala and frontal regions as a putative risk marker for depression. In line with previous studies (11-12,14,83-84), MDD patients showed decreased effective connectivity from left dorsolateral prefrontal cortex to left amygdala compared to controls. However, relatives did not differ from either controls or patients given the strong evidence (posterior probability > 0.95). Although evidence from separate studies support that both MDD patients and individuals at familial risk for MDD exhibit altered amygdala connectivity with the prefrontal cortex, the precise location of the coupling region within the prefrontal cortex may differ according to the groups. Since our DCM included only lateral prefrontal regions (BA 46 and 47) due to their prominent activation during the face-matching task, other prefrontal regions which are not included in the model, such as medial prefrontal regions (18,28), may play a more important role in risk for MDD.

### Resilience

During the face-matching task, relatives had higher effective connectivity from anterior cingulate cortex to left dorsolateral prefrontal cortex than controls and patients. Similar to the present findings, increased functional connectivity between the anterior cingulate cortex and left superior frontal gyrus in first-degree relatives was reported in our previous study as a marker of resilience (28). The anterior cingulate cortex is responsible for various aspects of emotional processing (85-86) and constitutes the cognitive-control network together with the dorsolateral prefrontal cortex (39). Previous studies have linked the neural coupling between the anterior cingulate cortex and the dorsolateral prefrontal cortex to a better performance in attention shifting (87) and increased top-down attentional control (88) in healthy participants. Thus, enhanced fronto-cingulate connectivity in relatives can serve as a resilience capacity mechanism by providing more cognitive control during affective face processing.

Interestingly, enhanced connectivity from the anterior cingulate cortex to dorsolateral prefrontal cortex was related to increased negative affect in MDD patients. Although previous studies associated enhanced fronto-cingulate connectivity with positive outcomes in healthy participants, this connection may have an altered association in MDD. Indeed, a recent study showed that weaker connectivity between anterior cingulate cortex and dorsolateral prefrontal cortex was beneficial for depression recovery in MDD patients before and after eight weeks of antidepressant treatment (89).

Furthermore, it is important to note that we here attempted to identify resilience-related changes in effective connectivity using a cross-sectional design. Although it brings some practical advantages, it is challenging to deem a unique feature as a resilience marker, first-degree relatives still have a chance of developing depression in future given the relatively wide range of age of onset of this disorder (90). Therefore, a unique alteration found in first-degree relatives can also reflect a risk marker that occurs before the onset of disease (28).

### Limitations

Our findings need to be interpreted in the light of some limitations. First, we assume that our experimental design measure neural responses to implicit emotional processing since all faces embodied expressions of emotion. However, the utilization of geometrical shapes in the control condition instead of neutral faces made it impossible to dissociate the effect of emotional valance from basic face processing. Thus, we could not investigate the direct effect of the emotional valance on brain activity and connectivity in this study.

Second, the current concept of resilience (20) requires that measures of stressor-load should be put into relation with symptomatology across time to assess resilience as an outcome. Therefore, the potential protective mechanisms found in our group comparisons might be of compensatory nature and their relevance for resilient stress-coping needs to be further evaluated.

Lastly, we here used DCM as a causal search paradigm by specifying a fully-connected model. Connection parameters which did not contribute to model evidence were then pruned away from the model. This procedure uses a greedy search algorithm which finds the best solution at each step rather than comparing all possible nested models. When the number of ROIs in the model increased, model space grows exponentially, and using a greedy search algorithm with large model space can introduce bias to construct post-hoc explanations for the surviving parameters (91).

## Conclusion

Our results suggest that the depressive state alters top-down control of higher visual regions, which are important for emotional face processing, whereas enhanced connectivity within the cognitive control network facilitates the capacity for resilience or compensation for familial risk factors. If further research, particularly longitudinal studies, supports these hypotheses, brain regions whose activity and connectivity pattern are related to the depressive state, risk and resilience can be used as targets in novel treatments of depression, such as neurofeedback (92-95).

## Data Availability

The data of the current study are available from the corresponding author,[C.W],upon reasonable request.

## Acknowledgements

The authors would like to thank Prof. Karl Friston for his valuable suggestions on the application of DCM and Dr. Peter Zeidman for his assistance with DCM analysis.

